# The Persistent Mortality and Heart Failure Burden of Anterior ST-Segment Elevation Myocardial Infarction Following Primary Percutaneous Coronary Intervention

**DOI:** 10.1101/2022.10.07.22280858

**Authors:** Jack L. Martin

## Abstract

**Background:** There is limited data on temporal trends in clinical outcomes after ST-segment elevation myocardial infarction (STEMI) treated with primary percutaneous coronary intervention (PCI) particularly beyond one year and in real world populations that include patients often excluded from randomized trials.

**Objectives:** We sought to compare the temporal trends in the incidence of death and re-hospitalization for congestive heart failure (CHF) following anterior STEMI in a Medicare cohort of beneficiaries treated with primary PCI in 2005 (n = 1,479) with those treated in 2016 through quarter (Q) 2 of 2017 (n = 22,432).

**Methods:** Outcomes were examined using both descriptive and regression analysis to control for differences in patient clinical characteristics over time.

**Results:** The 1-year mortality rate trended higher in the late cohort (10.3 vs 8.9%, p=0.068). The 2-year mortality rate was significantly higher in the late cohort (14.5 vs 11.4%, p<0.01). The one-year re-hospitalization for CHF was lower in the late cohort (10.6 versus 16.7%, p<0.01), but the 2-year rate was unchanged (19.3 vs 20.7%, p=0.55). After adjustment for covariates with two models there were highly statistically significant increases in mortality at 1-year (2.3 – 4.1%) and 2-years (4.2 – 6.5%) in the late cohort. The unadjusted trends in re-hospitalization for CHF persisted after adjustment for covariates.

**Conclusions:** Despite prior improvements in STEMI outcomes in the reperfusion era related to the broad adoption of timely PCI, there is a persistent high mortality and CHF burden in patients with anterior STEMI. New strategies that address reperfusion injury and enhance myocardial salvage are needed.

## Introduction

Ischemic heart disease remains the leading cause of death worldwide. Over fifteen million acute myocardial infarctions (MI) occur annually and forty percent of these are STEMIs requiring emergent reperfusion therapy. (1,2) The more widespread availability of timely reperfusion therapy, initially with thrombolysis and subsequently with primary percutaneous coronary intervention (PCI) resulted in a steady decline in post-MI mortality in many developed countries. (3-6) However, registry data suggest that there may be a plateau in these mortality improvements that was reached after national initiatives maximized access to primary PCI with stenting in developed markets. (4,5)

The observed improvements in clinical outcomes over time in all comers with STEMI could potentially be influenced by advancements in the PCI procedure or in secondary prevention measures in addition to the more widespread use of timely PCI. (4) Separating the impact of PCI utilization from the impact of these other potential advancements over time requires analysis of outcomes for comparable STEMI patients treated with primary PCI in different years. Published data on these temporal trends are limited, particularly with follow-up beyond one year and in real world primary PCI populations that include the higher risk patients often excluded from randomized trials. (7)

Several studies suggest that the incidence of congestive heart failure (CHF) post STEMI may have also decreased in the reperfusion era. However, these data come from relatively small studies and there are conflicting reports suggesting an increased incidence of CHF in patients surviving the acute STEMI event. (8-11) Further data are needed to address these crucial issues which relate to where continued efforts should be focused to mitigate the ongoing burden of STEMI. We thus sought to compare the temporal trends in the incidence of death and re-hospitalization for CHF following STEMI in Medicare beneficiaries treated with primary PCI in 2005 with those treated from 2016 to quarter (Q) 2 of 2017. Given the higher expected event rates in patients with anterior wall STEMI, the focus of this analysis is on this high-risk group. (9,12)

## Methods

The changes in one-year rates of mortality and re-hospitalization for CHF in patients treated for anterior STEMI in 2005 versus 2016 and Q1-Q2 of 2017 were examined using both descriptive and regression analysis. In order to allow for adequate follow-up, data were obtained from the 2005-2008 5% and the 2016-Q2 2019 100% Medicare Inpatient Limited Data Set populations. Patients discharged from an acute care hospital with a principal diagnosis of anterior STEMI were identified by ICD-9 diagnosis codes 410.00, 410.01, 410.10, 410.11 or ICD-10 diagnosis codes I21.01, I21.02, I21.09. Treatment with PCI with stenting was required and was identified in the earlier data by MS-DRGs 121, 122, 516, 526, 555, 557 and in the later data by MS-DRGs 246, 247. Patients with cardiogenic shock were excluded by eliminating patients with ICD-9 diagnosis code 785.51 and ICD-10 diagnosis code R57.0. This was done to avoid the potential confounding of the results by the inclusion of more shock patients in the late cohort.

The 2005-2007 Medicare Inpatient Limited Datasets only report the quarter of discharge from the hospital. Therefore, the 4-quarter as well as 8-quarter rates for mortality and re-hospitalization for CHF were assessed during this time frame and are reported as 1-year and 2-year results. The sample was divided into two cohorts based on the year of discharge from the hospital. The early cohort included cases discharged in 2005 and the late cohort included cases discharged in 2016 and in Q1-Q2 of 2017. The difference in outcomes between the early and late cohorts was assessed with t-tests. Regression analysis was also performed to control for differences in patient clinical characteristics over time. Medicare claims in 2005 reported only 9 secondary diagnoses whereas claims in 2016-2017 reported 24 secondary diagnoses. To ensure consistency in identifying comorbidities across the time frame, the first 9 secondary diagnoses were utilized in the 2016-2017 claims. The key independent variable in the regression analyses was an indicator for late cohort.

We estimated a linear regression of each outcome using the two risk adjustment strategies. In one set of regressions, propensity score matching (PSM) adjustment variables included age, sex, race/ethnicity, diabetes mellitus, hypertension, hyperlipidemia, atrial fibrillation and antioventricular block. In the second set of regressions, we controlled for Elixhauser comorbidities in addition to age, sex, and race/ethnicity. (13) In all regressions, we controlled for the quarter of hospitalization.

## Results

The baseline demographics and for the early (n =1,479) and late (n = 22,432) cohorts are outlined in table 1. The late cohort was younger and had fewer females and Caucasian patients compared with the early cohort. The late cohort had a higher Elixhauser Index with significantly more hypertension and hyperlipidemia. In addition, diabetes was more frequent in the late cohort with one year of follow-up. Atrioventricular block was less frequent in the later cohort. (table 1)

**Table 1.** Baseline Demographics.

Baseline Demographics
| Demographics |  | One Year Follow-up Group |  |  | Two Year Follow-up Group |  |  |  |
| --- | --- | --- | --- | --- | --- | --- | --- | --- |
| Patient Characteristic | Early Cohort<br>n = 1,479 | Late Cohort<br>n = 22,432 | Difference | p-value | Early Cohort<br>n = 798 | Late Cohort<br>n = 12,786 | Difference | p-value |
| Age (years) | 73.6±11.1 | 71.2±10.0 | -2.4 | 0.00 | 73.4±10.8 | 71.3±9.9 | -2.1 | 0.00 |
| Female | 48.8% | 34.7% | -14.1% | 0.00 | 50.6% | 35.0% | -15.6% | 0.00 |
| White | 88.6% | 86.7% | -1.9% | 0.03 | 88.2% | 87.4% | -0.8% | 0.48 |
| Black | 7.2% | 6.2% | -1.0% | 0.14 | 7.5% | 5.8% | -1.7% | 0.08 |
| Hispanic | 1.2% | 1.5% | 0.3% | 0.43 | 1.4% | 1.4% | 0.0% | 0.89 |
| Conditions |  |  |  |  |  |  |  |  |
| Diabetes | 23.9% | 27.6% | 3.7% | 0.00 | 26.1% | 27.5% | 1.4% | 0.37 |
| Hypertension | 60.8% | 70.3% | 9.5% | 0.00 | 61.0% | 69.8% | 8.8% | 0.00 |
| Hyperlipidemia | 44.6% | 58.0% | 13.4% | 0.00 | 43.2% | 58.6% | 15.4% | 0.00 |
| Atrial Fibrillation | 13.8% | 12.2% | -1.6% | 0.08 | 13.3% | 11.9% | -1.4% | 0.25 |
| AV Block | 5.6% | 2.5% | -3.1% | 0.00 | 6.6% | 2.4% | -4.2% | 0.00 |
| Elixhauser Index | 2.54 | 2.70 | 0.16 | 0.00 | 2.53 | 2.66 | 0.13 | 0.02 |

AV Block: atrioventricular block

Unadjusted outcomes are presented in table 2. Among Medicare beneficiaries diagnosed with anterior STEMI who are treated with PCI with stenting, the 1-year mortality rate trended higher in the late cohort at 10.3% versus 8.9% in the early cohort, p=0.068. The 2-year mortality rate was significantly higher in the late cohort at 14.5% versus 11.4% in the early cohort, p<0.01. One-year re-hospitalization for CHF was higher in the early cohort at 16.7% versus 10.6% in the late cohort, p<0.01, but the 2-year rate was unchanged (20.7% versus 19.3%, p=0.55).

**Table 2.** Rates of Mortality and Re-hospitalization for CHF by Year.

| Outcome | Early Cohort Mean | Later Cohort Mean | Unadjusted Difference | p-value |
| --- | --- | --- | --- | --- |
| 1-year mortality | 8.9% | 10.3% | 1.4% | 0.07 |
| 1-year re-hospitalization for CHF | 16.7% | 10.6% | -6.1% | 0.00 |
| 2-year mortality | 11.4% | 14.5% | 3.1% | 0.01 |
| 2-year re-hospitalization for CHF | 20.7% | 19.3% | -1.4% | 0.55 |

CHF: congestive heart failure

Risk-adjusted event rates are presented in table 3. After controlling for differences in clinical characteristics between earl and late cohorts using Elixhauser comorbidities there are highly statistically significant increases in the 1-year and 2-year mortality rates in the late cohort of 2.3 and 4.2 percentage points respectively. When controlling for differences in clinical characteristics using the PSM variables there are similar highly statistically significant increases in 1-year and 2-year morality rates in the late cohort of 4.1 and 6.5 percentage points. Risk-adjusted 1-year re-hospitalization rate for CHF decreased by 4.9 and 6.9 percentage points between the early and late cohorts with the PSM and Elixhauser risk adjustments respectively. However, there is no statistically significant change in the 2-year re-hospitalization rate for CHF between the early and late cohorts with either risk adjustment model.

**Table 3.**
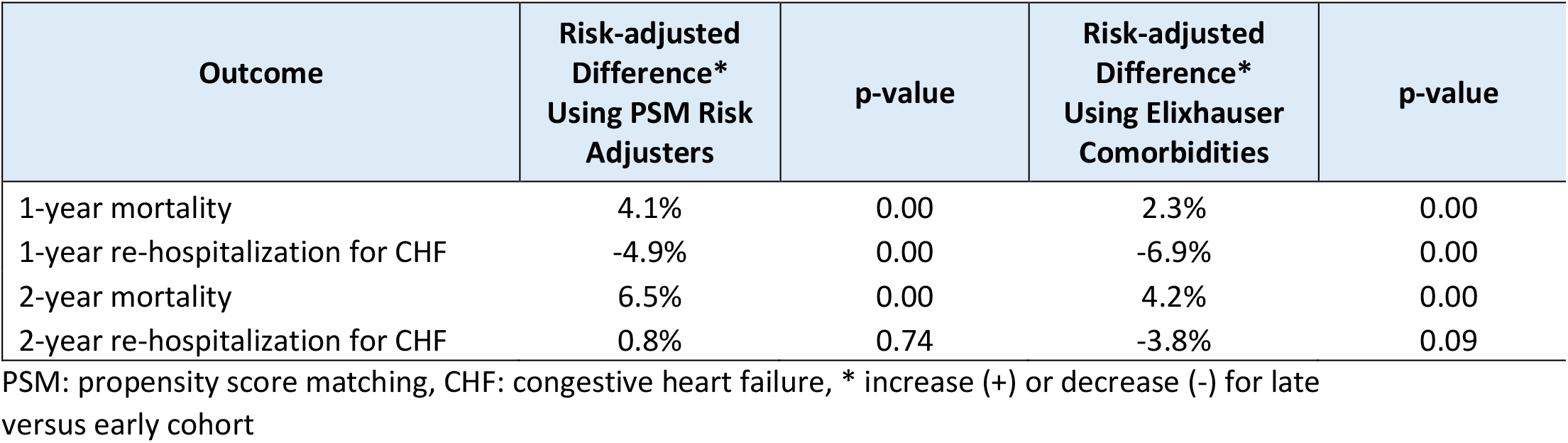
Risk-adjusted Difference in Outcome.

## Discussion

This analysis of outcome trends among Medicare beneficiaries who are diagnosed with anterior STEMI and treated with PCI yielded several important findings. Over time there was a trend for higher 1-year unadjusted mortality and a highly statistically significant increase in 2-year unadjusted mortality. Furthermore, the risk-adjusted 1-year and 2-year mortality rates increased significantly between the early and late cohorts when controlling for changes in clinical characteristics with the two different regression models. There was a significant decrease in the unadjusted and risk-adjusted 1-year re-hospitalization rates for CHF over time, but no statistically significant change in the 2-year re-hospitalization rates for CHF in either the unadjusted or risk-adjusted analyses.

The reduction in mortality for patients with STEMI in the reperfusion era is a major accomplishment of cardiovascular medicine. However, national registries suggest that there may not have been further progress after maximizing the benefit of early PCI in developed markets. (3-5) The rates of mortality and re-hospitalization for CHF in the present analysis support the premise that there has been no significant improvement in the last decade for STEMI outcomes when considering patients that received timely primary PCI with stenting. There are limited prior data focusing on these event rates over time only in patients treated with primary PCI for anterior STEMI. This is particularly the case for data on all-comers including patients not enrolled in randomized trials. (7, 14) A recent systematic review examining trends in PCI outcomes from 25 all-comers trials excluded STEMI patients. (15)

The present analysis not only finds no improvement in mortality over time in the anterior STEMI population treated with timely PCI but suggests a potential worsening. Given that these findings are demonstrated with two different risk-adjustment models, it is not likely that this is attributable to applying primary PCI to higher risk patients over time. In this regard patients with cardiogenic shock were excluded from the present analysis to control for the potential of more frequent triage of these high-risk patients to primary PCI over time and for the variable standard of care practices particularly related to the timing and choice of device escalation. The more widespread adoption of mechanical circulatory support in recent years may well translate into improved outcomes for this high-risk subgroup of STEMI patients experiencing cardiogenic shock. (16) Nonetheless, the present data suggests that there is a continued high mortality burden in Medicare beneficiaries with anterior STEMI treated with primary PCI in contemporary practice as evidenced by 1-year and 2-year death rates of 10.3% and 14.5% in the late cohort even after excluding patients with cardiogenic shock.

The explanation is not clear for the apparent improvement over time in the 1-year re-hospitalization rates for CHF which did not persist after longer follow-up. More effective use of standard of care medications for CHF may account for these improved outcomes, but that effect would be expected to be maintained at the 2-year analysis. It should be noted that during the follow-up of the late cohort there were payment policy changes related to early re-hospitalization for CHF which may have affected hospital coding. (17,18) These payment policy changes were followed by a well-documented decrease in per capita admissions for CHF in the United States after 2014. (19) Whether coding issues or an increased use of outpatient treatments for worsening heart failure accounts for these findings is not settled. (19) Regardless of the explanation for these findings, there is clearly a high persistent burden of CHF in Medicare beneficiaries with anterior STEMI treated with PCI based on the 1-year and 2-year re-hospitalization rates for CHF of 10.3% and 19.6% respectively in the late cohort analyzed in this study. These findings are consistent with a recent report on trends in post MI readmission for CHF in Medicare beneficiaries which noted a rate of 10.4% at one year in a less high-risk group not limited to only anterior STEMI. (20) These findings are also consistent with an analysis of temporal trends in Medicare beneficiaries demonstrating a significant increase in CHF incidence between 2011 and 2016 in patients with prior MI in contrast to a decreased incidence in patients with other comorbidities. (21)

CHF affects over 6 million Americans and is the leading cause of hospitalization in older adults. These patients have a nearly 50 percent 5-year mortality. (19, 22) The direct and indirect economic burden for CHF is over $100 billion globally and over $30 billion in the US alone. The recently completed PARADISE-HF trial demonstrated no additional benefit of sacubitril–valsartan compared with ramipril for the prevention of heart failure post MI and confirms the ongoing burden in a population treated predominantly with PCI for STEMI. (23) Given the increasing prevalence of CHF in an aging population the US economic burden related to CHF is projected to exceed $60 billion by 2030. (19,24) Ongoing efforts are clearly needed to track the burden of CHF specifically in survivors of STEMI and the impact of new reperfusion strategies on outcomes.

From 2005 to 2019 there have been advances in drug eluting stent technologies and in anticoagulant, antiplatelet and other pharmacologic therapies for acute and convalescent STEMI treatment. For example, drug eluting stents have been demonstrated to reduce re-hospitalization for restenosis in STEMI patients. (25) This reduction in restenosis may be associated with reduced mortality five years after STEMI, but not at earlier time points. (26) These devices and current pharmacologic therapies designed to optimize epicardial vessel patency do not address the reperfusion injury which may account for half of the residual infarct size after STEMI. (27-29) Infarct size is strongly associated with mortality and re-hospitalization for CHF after STEMI highlighting the importance of developing and implementing effective therapies to mitigate reperfusion injury. (30)

The lack of adjunctive therapies to primary PCI to further reduce myocardial damage post STEMI may offer an explanation for the presently observed persistence of high mortality and CHF re-hospitalization rates. Many pharmaceutical and device strategies have attempted to reduce reperfusion injury and infarct size. Despite positive results in preclinical models, these modalities have been largely ineffective in clinical trials. (28,31,32) Supersaturated oxygen (SSO2) therapy is the only adjunct device or drug demonstrated in an adequately powered randomized trial to reduce infarct size in STEMI patients treated with primary PCI and stenting. (33) A recent review details the effects of this therapy at the myocardial cell and microvascular level. (32) SSO2 therapy is FDA approved for intracoronary delivery after left anterior descending artery stenting for STEMI and is currently undergoing further investigation for expanded indications including in the unmet need of cardiogenic shock. Other modalities to address reperfusion injury including mechanical unloading, cooling therapies, and the delivery of other novel pharmaceutical agents require further study. (32) The broader adoption of effective adjunctive myocardial salvage therapies may offer additional benefit in reducing mortality and the incidence of CHF in STEMI populations.

## Limitations

The changes in the baseline demographics between the early and later cohorts were expected, but necessitated risk adjustment. The decrease in the proportion of female and Caucasian patients in the later cohort is consistent with other reports on trends in the demographics of Medicare patients with myocardial infarction. (34,35) The increase in comorbidities observed in the later cohort is also consistent with these prior reports. The decrease in the proportion of female patients likely accounts for the somewhat younger age of the late cohort given the known gender differences in the age of onset of ischemic heart disease. Changes in Medicare Inpatient Limited Datasets related to the number of secondary diagnoses reported over time could have affected the risk adjustment approach in ways that are not readily apparent. Nonetheless based on both the presently reported unadjusted and risk-adjusted event rates it is clear that mortality and re-hospitalization for CHF following primary PCI for anterior STEMI have not improved significantly in recent years.

The findings of the present study are limited to Medicare beneficiaries and further investigation is required to address outcomes in younger populations. The present analysis was undertaken in anterior STEMI patients because of the known higher event rates in this population which would maximize the ability to detect potential improvements in outcomes over time. It is unlikely that improvement in outcomes would be detected in the non-anterior STEMI population. We noted that STEMI patients with cardiogenic shock were excluded from the present analysis to avoid potential bias related to a higher proportion of these patients undergoing primary PCI over time, and confounding factors such as variability in the timing and prevalence of mechanical circulatory support for these patients. The present data cannot exclude the potential for primary PCI with mechanical circulatory support to improve outcomes in this high-risk subgroup over time.

## Conclusion

Despite prior improvements in STEMI outcomes in the reperfusion era related to the broad adoption of timely PCI, there is a persistent high mortality and CHF burden in patients with anterior STEMI. Improvement in outcomes beyond those achievable with restoring epicardial vessel patency will require implementation of strategies that enhance myocardial salvage by addressing downstream microvascular dysfunction and reperfusion injury.

## Data Availability

All data produced in the present study are available upon reasonable request to the authors

## Acknowledgement

The author acknowledges KNG Health Consulting for their assistance in data analysis.

## Notes

**Disclosure statement:** Zoll Medical Corporation funded the data analysis for the preparation of this manuscript. There are no other potential conflicts to disclose.

### Competing Interest Statement

Zoll Medical Corporation funded the data analysis for the preparation of this manuscript. There are no other potential conflicts to disclose.

### Funding Statement

Zoll Medical Corporation funded the data analysis for the preparation of this manuscript.

## References

1) GBD 2015 Disease and Injury Incidence and Prevalence Collaborators. Global, regional, and national incidence, prevalence, and years lived with disability for 310 diseases and injuries, 1990–2015: a systematic analysis for the Global Burden of Disease Study 2015. Lancet 2016; 388: 1545–602.

2) Chi GC, Kanter MH, Li BH, et al. Trends in Acute Myocardial Infarction by Race and Ethnicity. (J Am Heart Assoc. 2020;9:e013542.

3) Puymirat E, Simon T, Cayla G, et al. Acute myocardial infarction changes in patient characteristics, management, and 6-month outcomes over a period of 20 years in the FAST-MI Program (French Registry of Acute ST-Elevation or Non-ST-Elevation Myocardial Infarction) 1995 to 2015. Circulation. 2017;136:1908–1919.

4) Luscher TF, Obeid S. From Eisenhower’s heart attack to modern management: a true success story! Eur Heart J 2017; 38:3066–3069.

5) Szummer K, Wallentin L, Lindhagen L, et al. Improved outcomes in patients with ST-elevation myocardial infarction during the last 20 years are related to implementation of evidence-based treatments: experiences from the SWEDEHEART registry 1995–2014. Eur Heart J 2017;38: 3056–3065.

6) Garcia-Garcia C, Oliveras T, Serra J, et al. Trends in short- and long-term ST-segment-elevation myocardial infarction prognosis over 3 decades: a Mediterranean population-based ST-segment-elevation myocardial infarction registry. J Am Heart Assoc 2020;9:e017159.

7) Hosseiny AD, Moloi S, Chandrasekhar J, Farshid A. Mortality pattern and cause of death in a long-term follow-up of patients with STEMI treated with primary PCI. Open Heart 2016;3:e000405.

8) Cahill TJ, Kharbanda RK. Heart failure after myocardial infarction in the era of primary percutaneous coronary intervention: Mechanisms, incidence and identification of patients at risk. World J Cardiol 2017 May 26; 9(5): 407–415.

9) Hausenloy DJ, Garcia-Dorado D, Bøtkert HE, et al. Novel targets and future strategies for acute cardioprotection: Position Paper of the European Society of Cardiology Working Group on Cellular Biology of the Heart. Cardiovascular Research 2017; 113:564–585.

10) Moran AE, Forouzanfar MH, Roth GA, et al. The global burden of ischemic heart disease in 1990 and 2010: the Global Burden of Disease 2010 study. Circulation 2014;129:1493–1501.

11) Roger VL, Weston SA, Gerber Y, et al. Trends in incidence, severity, and outcome of hospitalized myocardial infarction. Circulation 2010;121:863–869.

12) Cung TT, Morel O, Cayla G, et al. Cyclosporine before PCI in patients with acute myocardial infarction. N Engl J Med 2015;373:1021–1031.

13) Elixhauser A, Steiner C, Harris DR, Coffey RM. Comorbidity measures for use with administrative data. Med Care 1998;36:8–27.

14) Virani SS, Alonso A, Aparicio HJ, et al. Heart disease and stroke statistics 2021 update: a report from the American Heart Association. Circulation. 2021;143:e254–e743.

15) Asano T, Ono M, Dai Z, et al. Temporal trends in outcomes after percutaneous coronary intervention: a systematic review of 66,327 patients from 25 all-comers trials. EuroIntervention 2021;17:online publish-ahead-of-print.

16) Helgestad OKL, Josiassen J, Hassager C, et al. Contemporary trends in use of mechanical circulatory support in patients with acute MI and cardiogenic shock. Open Heart 2020;7:e001214.

17) Daeho K, Makineni R, Panagiotou OA, et al. Assessment of Completeness of Hospital Readmission Rates Reported in Medicare Advantage Contracts’ Healthcare Effectiveness Data and Information Set. JAMA Network Open. 2020;3(4):e203555.

18) Gerhardt G, Yemane A, Apostle K, et al. Evaluating whether changes in utilization of hospital outpatient services contributed to lower Medicare readmission rate. Medicare & Medicaid Research Review 2014; 4(1):E1-E13.

19) Heidenreich PA, Fonarow GC, Opsha Y, etal. Economic issues in heart failure in the United States. Journal of Cardiac Failure (2022), doi:https://doi.org/10.1016/j.cardfail.2021.12.017.

20) Kochar A, Doll Ja, Liang L, et al. Temporal trends in post myocardial infarction heart failure and outcomes among older adults. J Card Fail 2021 Sep 11;S1071-9164(21)00361–4.

21) Khera R, Nitin Kondamudi N, Zhong L, et al. Temporal trends in heart failure incidence among medicare beneficiaries across risk factor strata, 2011 to 2016. JAMA Netw Open 2020;3(10):e2022190.

22) aylor CJ, Ryan R, Nichols L, Gale N, etal. Survival following a diagnosis of heart failure in primary care. Fam Pract. 2017 Apr 1;34(2):161–168.

23) Pfeffer MA, Claggett B, Lewis EF, et al. Angiotensin receptor–neprilysin inhibition in acute myocardial infarction. N Engl J Med 385;20:1845–1855.

24) Jackson SL, Tong X, King RJ, et al. Burden of Heart Failure Events in the United States, 2006 to 2014. Circ Heart Fail. 2018;11:e004873.

25) Van Leeuwen MAH, Daemen J, van Meighem NM, et al. Comparison of long-term outcomes of STEMI and NSTE-ACS after coronary stent placement: an analysis in a real world BMS and DES population. Int J Cardio 2013; 167:2082–2087.

26) Sabate M, Brugaletta S, Cequier A, et al. Clinical outcomes in patients with ST-segment elevation myocardial infarction with everolimus-eluting stents versus bare-metal stents (EXAMINATION): 5-year results of a randomized trial. Lancer 2016; 387:357–366.

27) Bulluck H, Yellon DM, Hausenloy DJ. Reducing myocardial infarct size: challenges and future opportunities. Heart 2016;102:341–348.

28) Heusch G, Gersh BJ. The pathophysiology of acute myocardial infarction and strategies of protection beyond reperfusion: a continual challenge. Eur Heart J 2017; 38:774–784.

29) Yellon DM, Hausenloy DJ. Myocardial reperfusion injury. N Engl J Med 2007;357:1121–35.

30) Stone GW, Selker HP, Thiele H, et al. Relationship between infarct size and outcomes following primary PCI: patient-level analysis from 10 randomized trials. J Am Coll Cardiol. 2016;67:1674–1683.

31) Davidson SM, Ferdinandy P, Andreadou I, et al. Multitarget strategies to reduce myocardial ischemia/reperfusion injury. J Am Coll Cardio 2019; 73:89–99.

32) Kloner RA, Creech JL, Stone GW, et al. Update for cardioprotective strategies for STEMI. J Am Coll Cardiol Basic Trans Science 2021. doi:10.1016/j.jacbts.2021.07.011.

33) Stone GW, Martin JL, de Boer MJ, et al. Effect of supersaturated oxygen delivery on infarct size after percutaneous coronary intervention in acute myocardial infarction. Circ Cardiovasc Interv. 2009;2:366–375.

34) Sack NC, Ash AS, Ghosh K. Recent national trends in acute myocardial infarction hospitalizations in medicare. Epidemiology 2015;26(4).

35) Krumholtz HM, Normand ST, Wang Y, et al. Twenty-year trends in outcomes for older adults with acute myocardial infarction in the united states. JAMA Netw Open 2019;2(3): e191938.

